# Food Insecurity in Households of People with Autism Spectrum Disorder during the COVID-19 Pandemic

**DOI:** 10.1101/2021.03.30.21254339

**Authors:** Vijay Vasudevan, Arun Karpur, Andy Shih, Thomas Frazier

## Abstract

**Objectives:** To explore differences in food insecurity for individuals and families of people with autism spectrum disorder (ASD) during the COVID-19 pandemic by individual, family, and neighborhood characteristics.

**Methods:** We surveyed a convenience sample of households of people with ASD. We calculated food insecurity using items from the US Census Bureau’s Household Pulse Survey..

**Results:** Over half of all respondents reported being food insecure (51.8%). Respondents who reported being food insecure were more likely to be minority, have a high school education or less, be on public insurance or uninsured, live in urban/rural communities, and say that their community is not supportive. The majority of respondents did not get free food or groceries (53.2%). Food insecure respondents who got free food was most likely to get them from schools (34.2%).

**Conclusion:** This is the first study of its kind to explore food security in households of people with ASD. The pandemic has exacerbated existing neighborhood disparities. The federal response to food insecurity caused by the pandemic needs to be further explored especially for preferred and medically necessary foods for people with ASD.

## INTRODUCTION

The US Department of Agriculture (USDA) defines food insecurity when a household’s “access to adequate food is limited by a lack of money and other resources.”^1^ Prior to the coronavirus disease 2019 (COVID-19) pandemic, 13.6% of all households with children were food insecure, with 6.5% of children facing food insecurity (∼2.4 million households).^1^ People with disabilities and households of individuals with autism spectrum disorder (ASD) were at a greater risk for food insecurity when compared to the general population.^2,3^ Studies have demonstrated the short- and long-term physical and mental health impacts of food insecurity.^4^ According to the US Census Bureau’s Household Pulse Survey (which asks how the coronavirus pandemic is impacting households), 18.3% of households with children experienced food insufficiency in the last seven days.^5^

The coronavirus pandemic has greatly exacerbated food insecurity.^6^ Leddy et al. illustrated how the COVID-19 pandemic (1) built on pre-existing disparities, (2) introduced its own stressors, (3) how food insecurity impacted household stress, behavioral, and inflammatory pathways, (4) impacted the physical and mental health outcomes, and (5) had a feedback loop back to the previous levels.

It is unclear how many households of individuals with ASD are included in the current food insecurity estimates during the COVID-19 pandemic. Therefore, the purpose of this study is to provide an initial estimate for food insecurity for households with someone with ASD by different individual, family, and neighborhood characteristics.

## METHODS

This study was approved as exempt by the Institutional Review Board at [insert name of author’s IRB]. This study used a convenience national sample, anonymous online survey from November 18, 2020 through December 7, 2020.

Food insecurity items were modified from the Household Pulse Survey specifically for households with someone with ASD.^5^ We then coded the responses into food secure, food insecure, and very low food security based on the USDA’s definitions.^1^ We modified the items to ask if anyone in the household received free food or groceries from a list of seven options such as school, church, or food bank. We asked about approximate annual income and family size to estimate Federal Poverty Level (FPL). Neighborhood quality items were modified from the National Survey of Children’s Health.^7^ Self-reported physical and mental health items were modified from the Behavioral Risk Factor Surveillance System.^8^

We collapsed food insecurity into a dichotomous variable where food insecure and very low food security being coded as food insecure. Frequency and chi-square analyses were performed to explore differences in food security status. Significance was set at alpha equal to 0.05 a priori. Frequency of where food insecure respondents for free groceries or meals were performed.

## RESULTS

This study had respondents (n=1515) from 48 states and the District of Columbia (excluding Wyoming and South Dakota) with over 21% of the respondents from New York and California. Parents and guardians of people with ASD accounted for a majority of the respondents (89.2%), with autistic adults accounting for the remainder of respondents. The majority of respondents were between 35 and 54 years old (57.1%), female (85.4%), White, non-Hispanic (65.0%), had more than a high school education (65.1%), had greater than 200% FPL (55.9%), lived in a suburban environment (52.9%), and do not live in a supportive neighborhood (89.1%). Nearly two-thirds of all respondents (62.4%) reported that the COVID-19 pandemic impacted their family “a lot.” Finally, over half of all respondents reported some food insecurity (19.5% reported being “food insecure” and 32.3% reported “very low food security”).

There was no significant difference for food insecurity by Census region. The region with the highest rate of food insecurity was the South (34.8%) followed by the Northeast (24.1%), West (23.1%) and Midwest (17.9%). Table 1 presents the comparison of respondents reporting being “food secure” compared to either “food insecure.” Respondent gender was the only variable which did not significantly associate with food security status.

**Table 1:** Food Insecurity Status by Individual and Neighborhood Characteristics.

|  | Food secure | Food insecure |
| --- | --- | --- |
| Age of respondent*** |  |  |
| Less than 45 years old | 40.2% | 59.8% |
| 45 years old or older | 59.2% | 40.8% |
| Gender |  |  |
| Female | 47.8% | 52.2% |
| Male | 51.8% | 48.2% |
| Race*** |  |  |
| White, non-Hispanic | 56.2% | 43.8% |
| Black, non-Hispanic | 34.3% | 65.7% |
| Hispanic | 25.5% | 74.5% |
| Education*** |  |  |
| High school graduate or less | 28.8% | 71.2% |
| More than high school | 58.6% | 41.4% |
| Federal Poverty Level*** |  |  |
| Less than or Equal to 200% FPL | 20.0% | 80.0% |
| Greater than 200% FPL | 64.1% | 35.9% |
| Received SNAP benefits*** |  |  |
| Did not receive SNAP | 60.1% | 39.9% |
| Received SNAP | 26.3% | 73.7% |
| Insurance coverage for child with ASD*** |  |  |
| Any Private (includes dual eligible) | 67.1% | 32.9% |
| Public only (Medicaid/Medicare) | 30.1% | 69.9% |
| Uninsured | 32.5% | 67.5% |
| Level of support for child with ASD*** |  |  |
| A little to no support | 56.5% | 43.5% |
| A medium amount of support | 50.9% | 49.1% |
| A lot of support | 41.3% | 58.7% |
| Respondent self-reported physical health*** |  |  |
| Excellent, Very good, or Good | 54.6% | 45.4% |
| Fair, or Poor | 34.8% | 65.2% |
| Respondent self-reported mental health*** |  |  |
| Excellent, Very good, or Good | 56.5% | 43.5% |
| Fair, or Poor | 41.5% | 58.5% |
| Neighborhood*** |  |  |
| Urban | 35.1% | 64.9% |
| Suburban | 58.7% | 41.3% |
| Rural | 37.9% | 62.1% |
| Neighborhood supportiveness** |  |  |
| Lives in supportive neighborhood | 59.6% | 40.4% |
| Does not live-in supportive neighborhood | 46.7% | 53.3% |
| Level of coronavirus pandemic impact*** |  |  |
| Not at all, Very little, or Somewhat | 57.4% | 42.6% |
| A lot | 42.7% | 57.3% |
| ** p < 0.01, *** p < 0.001 |  |  |

A majority of respondents who were food insecure did not receive any free groceries or free meals (53.2%). Respondents who were food insecure who received free meals or groceries got them from schools (34.2%), followed by a food bank or pantry (25.6%), family/friends (21.8%), community program (15.2%), or church/synagogue/religious institution (12.2%).

## DISCUSSION

This is the first known study of its kind to assess food insecurity from a sample of households with people of ASD. The major finding was that the COVID-19 pandemic increased food insecurity for families with ASD and makes them vulnerable to physical and mental health impacts. Food insecurity during the COVID-19 pandemic was impacted at the individual-, family-, and neighborhood-level.

Since the pandemic, the United States implemented a mixture of federal responses to address the economic disparities and subsequent food insecurity.^9^ Despite the federal response, food insecurity is a still a threat to the country. President Biden recently signed an Executive Order expanding food assistance programs like SNAP and the Pandemic Electronic Benefits Transfer for children who are missing meals due to school closures.^10^ During the pandemic, schools offered a combination of in-person and online education, therefore it is imperative to understand how students with ASD and their families receive food during this period including preferred foods and accommodations for necessary allergies.^11^ Subsequently, when the school are closed, for pandemic reasons or the students need to quarantine, researchers and policy makers need to create safety nets for families of people with ASD to make sure that the students and their families have access to the preferred and medically necessary food. Future studies need to explore how these policy and future initiatives impacts food insecurity for families of people with ASD.

When compared to Census geographic regions^1^, families of people with ASD reported more food insecurity during the coronavirus pandemic regardless of region. The pandemic has made the food system within the United States even more vulnerable for families of people with ASD. With food deserts and their accompanying health risks being well established and well-known,^12^ this study revealed that a greater percentage of families of people with ASD living in urban or rural environments were more likely to report being food insecure than people living in suburban communities. Additionally, respondents who reported living in neighborhoods that were not supportive were more likely to report being food insecure than families being food secure. One aspect that needs to be further explored is what role Race/Ethnicity plays at the intersection of neighborhood, food environments, and food insecurity during the pandemic.

This study has two primary limitations. Firstly, the study used a convenience sample of families of people with ASD and was not representative of households of people with ASD. The second limitation was that this study did not use the full 18 USDA food insecurity item set. Despite these limitations, this study establishes a baseline estimate of food insecurity for households of people with ASD during the COVID-19 pandemic.

### Public Health Implications

Food insecurity has long term ramifications and the COVID-19 pandemic has made its effects much more prevalent especially for households of people with ASD. For people with ASD and their families, food insecurity could have additional impact on their physical and mental health status. Policy makers should consider strategies to improving access to preferred and medically necessary food. Additional efforts should be undertaken to improve community design especially at the intersection of neighborhood, race, and disability status.

## Data Availability

Data for this study will not be made publicly available.

## REFERENCES

1. Coleman-Jensen A, Rabbitt MP, Gregory CA, Singh A. Household Food Security in the United States in 2019. U.S. Department of Agriculture, Economic Research Service; 2020:47. https://ers.usda.gov/webdocs/publications/99282/err-275.pdf?v=9716.7

2. Heflin CM, Altman CE, Rodriguez LL. Food insecurity and disability in the United States. Disabil Health J. 2019;12(2):220–226. doi:10.1016/j.dhjo.2018.09.006

3. Karpur A, Vasudevan V, Frazier T, Shih A, Lello A. Food Insecurity in the Households of Children with Autism Spectrum Disorders and Intellectual Disabilities in the U.S.: Analysis of the National Survey of Children’s Health Data 2016 - 2018. Autism. (in publication).

4. Gunderson C, Seligman HK. Food Insecurity and Health Outcomes. Econ Voice. 2017;14(1):0–0. doi:10.1515/ev-2017-0004

5. US Census Bureau. Measuring Household Experiences during the Coronavirus Pandemic. Census.gov. Accessed January 20, 2021. https://www.census.gov/householdpulsedata

6. Leddy AM, Weiser SD, Palar K, Seligman H. A conceptual model for understanding the rapid COVID-19–related increase in food insecurity and its impact on health and healthcare. Am J Clin Nutr. 2020;112(5):1162–1169. doi:10.1093/ajcn/nqaa226

7. National Survey of Children’s Health. Data Resource Center for Child and Adolescent Health. Accessed January 20, 2021. https://www.childhealthdata.org/learn-about-the-nsch/NSCH

8. Centers for Disease Control and Prevention. Behavioral Risk Factor Surveillance System. Published August 31, 2020. Accessed January 26, 2021. https://www.cdc.gov/brfss/index.html

9. Kinsey EW, Kinsey D, Rundle AG. COVID-19 and Food Insecurity: an Uneven Patchwork of Responses. J Urban Health. 2020;97(3):332–335. doi:10.1007/s11524-020-00455-5

10. Fact Sheet: President Biden’s New Executive Actions Deliver Economic Relief for American Families and Businesses Amid the COVID-19 Crises. The White House. Published January 22, 2021. Accessed January 27, 2021. https://www.whitehouse.gov/briefing-room/statements-releases/2021/01/22/fact-sheet-president-bidens-new-executive-actions-deliver-economic-relief-for-american-families-and-businesses-amid-the-covid-19-crises/

11. Cermak SA, Curtin C, Bandini LG. Food selectivity and sensory sensitivity in children with autism spectrum disorders. J Am Diet Assoc. 2010;110(2):238–246. doi:10.1016/j.jada.2009.10.032

12. Beaulac J, Kristjansson E, Cummins S. A Systematic Review of Food Deserts, 1966-2007. Prev Chronic Dis. 2009;6(3).

